# AI and the Eye – Integrating deep learning and *in silico* simulations to optimise diagnosis and treatment of wet macular degeneration

**DOI:** 10.1101/2024.02.13.23299445

**Authors:** Rémi J. Hernandez, Wahbi K. El-Bouri, Savita Madhusudhan, Yalin Zheng

**Author notes:** **Study Team** Chief Investigator: Prof Yalin Zheng, Co-investigators: Dr Wahbi El-Bouri, Dr Savita Madhusudhan, Mr Remi Hernandez Statistician: Dr Wahbi El-Bouri.

## Abstract

This protocol describes the A-EYE Study and provides information about procedures for entering participants. Every care was taken in its drafting, but corrections or amendments may be necessary. These will be circulated to investigators in the Study. Problems relating to this Study should be referred, in the first instance, to the Chief Investigator.

This study will adhere to the principles outlined in the UK Policy Framework for Health and Social Care Research (v3.2 10^th^ October 2017). It will be conducted in compliance with the protocol, the UK General Data Protection Regulation and Data Protection Act 2018, and other regulatory requirements as appropriate.

**DESIGN:** Single centre non-interventional study of patients with age-related macular degeneration to develop computational models of disease prediction and treatment outcome involving analysis of macular OCTA scans.

**AIMS:** *Primary Objective:*

- To develop a mathematical model (or *in silico* model) of blood flow and anti-VEGF transport in the retina that, in combination with AI-based analysis of macular OCTA scans and clinical data, can be used to predict treatment response in patients with neovascular age-related macular degeneration (nAMD).

*Secondary objectives:*

- To apply deep learning models in combination with *in silico* models of blood flow to OCTA analysis, to confirm diagnosis of nAMD and its clinical subtypes.
- To develop prognostic models to predict treatment outcome based on longitudinal patient follow-up.
- Using *in silico* simulations, to understand why certain patients do not respond optimally to anti-VEGF treatment.
- To define and simulate individualised anti-VEGF treatment for optimal response.

**OUTCOME MEASURES:** A validated *in silico* model of patient response to nAMD and anti-VEGF treatments tailored to individual patients using OCTA scans.

- Identify optimal intravitreal anti-VEGF therapy drug regime for individual patients using *in silico* models
- Improve on the classification and characterisation of neovascular AMD into its subtypes
- Predict risk factors for poor treatment outcomes such as retinal vascular topology

**POPULATION ELIGIBILITY:** All patients aged 55 years or more, with a new diagnosis of nAMD in at least one eye, attending the Macular Clinic at Royal Liverpool University Hospital, who have had a macular OCTA scan at baseline i.e. at the time of diagnosis.

**DURATION:** 48 months

*Clinical Queries:* Clinical queries should be directed to Dr Savita Madhusudhan who will re-direct the query to the appropriate person if necessary.

*Sponsor:* The University of Liverpool is the research Sponsor for this Study. For further information regarding the sponsorship conditions, please contact: Alex Astor Head of Research Support – Health and Life Sciences University of Liverpool Research Support Office 2nd Floor Block D Waterhouse Building 3 Brownlow Street Liverpool L69 3GL

*Funder:* EPSRC DTP in AI and Future Digital Health is funding the studentship associated with this study. Mr Remi Hernandez is the PhD candidate holding the studentship and Dr El-Bouri, Prof Zheng, and Dr Madhusudhan are his supervisors.

## 1. INTRODUCTION

### 1.1 BACKGROUND

Age related macular degeneration (AMD) accounts for 8.7% of blindness worldwide1, and is the leading cause of vision loss in people over the age of 60 in developed countries2. The underlying pathology in wet AMD (also known as neovascular AMD (nAMD)) is the development of abnormal neovascularisation in the central area of the retina, called the macula. The ophthalmic team at the Royal Liverpool University Hospital currently routinely use the gold standard method of diagnosing nAMD – fluorescein angiography (FA) alongside indocyanine green angiography (ICGA) and optical coherence tomography (OCT) scans of the macula, based on which nAMD is classified into subtypes. Dye-based retinal angiography (FA and ICGA), however, is invasive, time-consuming, costly, and has risks, including death from anaphylactic reactions to the dye rarely. After diagnosis, the main treatment method is often a prolonged course of VEGF inhibitor drug injected into the eye. However, up to one quarter of patients on anti-VEGF drugs do not respond optimally to treatment3. This may be due to various patient and disease characteristics, all which are not well understood.

Recently, optical coherence tomography angiography (OCTA) has emerged as a novel, quick, non-invasive imaging modality that provides detailed, high-resolution images of the retinal vasculature without the risks of FA. The recent success of AI and *in silico* simulations in various biological applications is expected to be key in realising the potential of OCTA to replace FA in nAMD diagnosis and management and may have a role to play in research and development of newer pharmacological interventions for nAMD in future.

### 1.2 RATIONALE FOR CURRENT STUDY

The prognosis for patients with nAMD has improved over the years due to the use of anti-VEGF treatments. These are long-term repeated intravitreal injections. Response to treatment has been defined as optimal (good), poor or none taking into consideration both morphological features on OCT and functional outcome in terms of visual acuity^4^ and multiple factors may account for this^3^. A further proportion of patients exhibit tachyphylaxis due to repeated administration of the treatment^5^. Therefore, there is an on-going need to understand variations in treatment-response and, furthermore, to determine whether individual dosing regimens can be optimised with available existing anti-VEGF therapy. To investigate this, *in silico* models of the retina will be developed based on patient OCTA scans. *In silico* models are computer simulations of the underlying maths/physics behind the biology. Recently, these models have been used to simulate blood flow^6^, oxygen transport in the retina^7^, and pharmacokinetics^8^ in eyes with and without AMD. This study will take OCTA scans of patients being treated with anti-VEGF agents in order to extract the geometric features of the retinal vasculature specifically delineating the MNV complex. These will be segmented, and blood flow and oxygen transport simulations will be conducted to determine potential regions of hypoxia/VEGF release.

These models will then be extended to simulate anti-VEGF treatments looking at optimal dose timings for individuals. The validation of the models will come from the longitudinal scans to see if the models can accurately predict nAMD response to treatment. Sub-optimal response can be investigated *in silico* by investigating the nature of the MNV, choice of treatment (e.g. combination of anti-VEGF therapy with photodynamic therapy vs. anti-VEGF monotherapy), varying treatment protocols (e.g. fixed dose, treat and extend) with anti-VEGF drug regimes, and most-importantly, helping to identify patients for future clinical trials for newer drugs by understanding variations in disease pathology and predict need and frequency of future monitoring after treatment completion.

Different subtypes of nAMD (types 1, 2 and 3 and aneurysmal type 1)^17,18^ are well characterised on conventional retinal imaging modalities of FA, ICGA and OCT, and although understanding of associated OCTA features is certainly growing, very little research has looked at diagnosing the main subtypes of nAMD using OCTA and AI, and exploring characteristics in relation to the subtype of MNV and visual outcomes with treatment. This is important as it predicts treatment burden^9^ and holds prognostic value^10^.

We will aim to use convolutional neural networks (CNNs) to confirm diagnosis of nAMD and its clinical subtype from OCTA scans. Recent work has looked at analysing OCTA features in response to anti-VEGF therapy in some subtypes of nAMD^10,11^. The high volume of data generated by 3-D macular OCTA scans, the need for manually correcting segmentation before analysing features, multiple quantitative metrics that are available for analysis, the impact of artifacts on accurate interpretation of the scans – all these make this new technology which is gaining traction rapidly in clinical practice, ideally suited for automated analysis and application of AI methods to enable computer-aided diagnosis and management of this visually debilitating condition.

## 2. STUDY OBJECTIVES

### Primary Objective

- To develop a mathematical model (or *in silico* model) of blood flow and anti-VEGF transport in the retina that, in combination with AI-based analysis of macular OCTA scans and clinical data, can be used to predict treatment response in patients with nAMD.

### Secondary objectives

- To apply deep learning models in combination with *in silico* models of blood flow to OCTA analysis, to confirm diagnosis of neovascular AMD and its clinical subtype.
- To develop prognostic models to predict treatment outcome based on longitudinal patient follow-up.
- Using *in silico* simulations, to understand why certain patients do not respond optimally to anti-VEGF treatment.
- To define and simulate individualized anti-VEGF treatment for optimal response.

## 3. STUDY DESIGN

This is a single centre data analysis project.

- Proposed study start date: 1^st^ December 2022
- Proposed study end date: 30^th^ November 2027
- Duration: 60 months

We aim to include 400 patients; data collection will be both retrospective and prospective. Primary endpoint time will be 3 years from the onset of treatment.

Patients over the age of 55 years with a new diagnosis of nAMD in at least one eye, attending the Macular Clinic at Royal Liverpool University Hospital, either for treatment or monitoring, who have had a macular OCTA scan at baseline will be eligible.. There is no study-related active intervention or deviation from standard clinical care as determined by the clinician, other than additional OCTA scans in the study eye at defined time points during follow-up (see below).

Data collected will include patient demographics, subtype of nAMD as diagnosed at baseline by clinicians using multimodal retinal imaging in the study eye(s), choice of intravitreal antiVEGF drug, duration of antiVEGF therapy, treatment regime including frequency of injections over a maximum of 3 years from baseline, best-recorded visual acuity at baseline, month 4 (i.e. 8 weeks after completion of the ‘loading phase’ of antiVEGF therapy which includes 3 four-weekly injections), months 12, 18, 24, 30 and 36 from baseline. For retrospective analysis, all available OCTA scans in the study eye(s) of included patients will be utilised; for the prospective arm of the study, OCTA scans will be acquired at baseline, month 4, end of year 1 and thereafter every 6 months for a maximum of 3 years from baseline, amounting to a total of 7 OCTA scans.

A minimum set of patient personal information will be collected including: NHS number, Initials, Name, Contact details, Age and gender. Age and gender will be required for the in silico computational modelling while the others will be used to check patient records, follow up patients and to make sure that the research is being done properly. This identifiable and sensitive data will only be accessible by the direct care team led by Dr. Savita Madhusudhan and the student for day-to-day management of the study.

### 3.1 STUDY OUTCOME MEASURES

A validated *in silico* model of patient response to nAMD and anti-VEGF treatments tailored to individual patients using OCTA scans.

## 4. PARTICIPANT ENTRY

### 4.1 PRE-REGISTRATION EVALUATIONS

Confirmation of a strong suspicion of or definite diagnosis of nAMD as determined by medical retina specialist(s) at the study site based on standard investigations in the departmental AMD pathway.

### 4.2 INCLUSION CRITERIA

Patients over the age of 55 years with a new diagnosis of nAMD in at least one eye, attending the Macular Clinic at Royal Liverpool University Hospital, either for treatment or monitoring, who have had one or more macular OCTA scans acquired as part of their routine clinical care including one at baseline before commencement of anti-VEGF treatment who are willing to take part in the study and are able and willing to give a valid consent.

### 4.3 EXCLUSION CRITERIA

Patients with peripapillary idiopathic polypoidal choroidal vasculopathy, previous retinal laser or antiVEGF intravitreal therapy, past history of posterior uveitis, other retinal conditions that might confound macular changes related to a diagnosis of nAMD including moderate or severe diabetic retinopathy, choroidal neovascularisation and macular oedema related to causes other than nAMD. Any underlying condition(s) that will prevent acquisition of good quality OCTA images.

### 4.4 WITHDRAWAL CRITERIA

The participant can withdraw from the study at any time, without explanation. He/she will be allowed to withdraw from the study and have the data retained or to withdraw from the study and have the data removed. For participants involved in the prospective study, their withdrawal will not impact any of their clinical care.

## 5. ADVERSE EVENTS

The study is non-interventional and will not involve any new procedures other than additional OCTA scans as specified above or any additional clinic visits; patients will otherwise receive standard care as part of the departmental AMD management guidelines under the discretion of the treating clinician. The research team do not expect there will be any adverse events (AE) or serious AE (SAE), however, in the event of any occurring, the procedures below will be followed. The CI of the study will report the reportable events to the sponsor immediately, but not later than 3 calendar days after investigational site study personnel’s awareness of the event.

## 6. ASSESSMENT AND FOLLOW-UP

All assessment and follow-up will be clinician determined as part of the on-going course of treatment or monitoring. Standard care involves any patient with a new diagnosis of nAMD being seen by a clinician in the Macular Clinic within 2 weeks of diagnosis, and in instances where a decision to treat is made, patients will be consented for a course of intravitreal antiVEGF drug therapy that involve a ‘loading phase’ of 3 injections given every 4 weeks and thereafter either i) a ‘fixed regime’ with 8 weekly injections till the end of year 1, followed by ‘treat and extend’ until the disease activity resolves or ii) an ‘early treat and extend’ where by injection interval is extended as per clinician discretion after the loading phase is complete, and continued until the disease activity has resolved. At baseline patients would have had retinal angiograms for confirmation of diagnosis, an OCT, OCTA, colour fundus and fundus autofluorescence images and at every follow-up visit, they receive an OCT scan, as part of the standard treatment pathway; additional retinal imaging at follow-up visits will be at clinician discretion. When treatment is deemed to be complete, patients are followed up for a further 2 years as part of the standard care as there is a risk of disease reactivation. Treatment and monitoring intervals, especially after the first year and length of follow-up is determined by individual patient response to therapy. For the purpose of this study, there is no other change to the patient pathway apart from the additional acquisition of follow-up OCTA scans at visits coinciding with visits after the ‘loading phase’, end of month 12, 18, 24, 30 and 36. This is acquired at the same time as acquiring the OCT scan, on the same machine and will only add another 2-3 minutes. Data to be collected: Diagnosis with respect to AMD subtype, patient demographics, visual acuity, details of treatment for AMD, retinal imaging for analysis, mainly OCTA from both treatment-naïve and treated patients, but other modalities of retinal imaging over multiple clinic visits, to be reviewed where available, and if felt necessary to understand and analyse findings on OCTA (please refer also to Study Design).

A standard questionnaire to gather information on self-reported vision-targeted health status will be administered at time points when a OCT-A is acquired on the prospective arm of the study, which will provide patient reported outcome measures to be included in our analyses. (More information is available at https://www.nei.nih.gov/learn-about-eye-health/resources-for-health-educators/outreach-materials/visual-function-questionnaire-25; the questionnaire is additionally submitted along with the study protocol)

## 7. STATISTICS AND DATA ANALYSIS

### 7.1 SAMPLE SIZE

As an observational analytic cohort study design, this trial requires no power calculation for estimates of effect. However, we will seek to recruit up to 400 patients including approx. 200 patients in the prospective arm of the study. For patients in whom retrospectively available OCTA scans are present, these will be included, in addition to up to 7 scans for prospectively recruited patients; this will be sufficient to develop novel AI algorithms and *in silico* models for the prediction of nAMD subtype and prognosis.

### 7.2 DATA ANONYMISATION

An encrypted file will be generated and each participant assigned a unique identification number (UID) in order to protect confidentiality and this UID index file will be stored on hospital’s network drive and only be accessed by the direct care team led by Dr Savita Madhusudhan, and the student already having an honorary contract with the Trust for the day-to-day management of the study. The clinical data will be collected from the existing electronic patient record database as well as images from Heidelberg (Heidelberg Engineering, Heidelberg, Germany) imaging software database with all identifying information removed and indexed by the UID at the time of anonymisation. All the anonymised images and clinical data will be stored on the hospital’s network drive for record.

### 7.3 DATA TRANSFER

The pseudo-anonymised data indexed by UID will be subjected to deep learning algorithmic interpretation at the University of Liverpool due to the high demand of GPU computing power. Pseudo-anonymised data (images and clinical data without any personal identifying information) will be transferred by using encrypted memory key to a password-protected University computer and stored on the secured University network drive. A data transfer agreement will be sought between the hospital Trust and the University before any data is transferred. Staff and researchers at the University of Liverpool (UoL) will only have access to the anonymised dataset for the analysis. No patient confidential information will be seen or accessed by the UoL researchers at any time. Any queries regarding the patient data will be dealt with by the members of the direct care team.

### 7.4 DATA ANALYSIS

During the study, all pseudo-anonymised patient information will be kept safe and secure in the University of Liverpool’s Active Data Store. The Active Data Store is a centralised secure data storage facility for electronic research data managed by the Liverpool Research Data team.

Anonymised OCTA images with clinical data collected from the participants will be used for the research on deep learning based prognostic methods. *The analysis will be performed by the research team at the University. Please note, this is a fundamental data-driven research that will study new machine learning ideas and methods with potentials for the prediction of disease progression. These new methods are not intended to be used for the management of patients within this project, there would be no any risk or safety concerns to patients. If these new methods are proved useful, further developments and clinical validation will be needed to make them as a medical device, but they are out of the scope of the study*.

Although we expect the develop models will use 2 to 5 sequential images to make the prediction, given the relatively small number of patients and the data hunger of deep learning, we will create more sequences by taking portions from longer sequences so as that we will have an ‘augmented’ dataset. The anonymised data will be split into training, validation and testing sets in a ratio of 70:20:10 for example. The training and validation datasets will be used to train AI modes while the testing set will be kept unseen during the model training and only be used for the evaluation of the performance of the trained models. Images of any single patient will only be used in either training or testing in order to avoid information leakage. All the images will be pre-processed so that the size and intensity values are compatible with deep learning models.

The primary outcome measure of diagnostic and prognostic accuracy for the AI and *in silico* modelswill be area under the curve (AUC), as well as the optimum sensitivity and specificity, based on Youden’s index^12^. We will use DeLong’s method^13^ to calculate the 95% confidence intervals and to test for a statistically significant difference between AUCs. For sensitivity and specificity, we will calculate bootstrapped 95% confidence intervals, with 2000 samples. The positive predictive values (PPV) and negative predictive value (NPV) will also be reported.

All the statistical analysis will be performed using statistical software Python 3 and/or R-studio as appropriate.

## 8. REGULATORY ISSUES

### 8.1 ETHICS APPROVAL

The Chief Investigator will obtain approval from the National Research Ethics Committee (NREC) and Health Research Authority (HRA) approval. The study will be conducted in accordance with the recommendations for physicians involved in research on human subjects adopted by the 18th World Medical Assembly, Helsinki 1964 and later revisions.

### 8.2 CONSENT

#### Retrospective arm

The UoL researchers will only use pseudo-anonymised data. As a retrospective study we will be accessing confidential patient information without consent in England. Therefore, ethical approval will be sought on the basis of health and social care research in the public interest through an application to the Confidentiality Advisory Group (CAG) using the HRA Integrated Research Application System (IRAS).

#### Prospective arm

A patient information leaflet explaining the objectives of this non-interventional study will be given to eligible patients attending the Macular Service at St. Paul’s Eye Unit, Royal Liverpool University Hospital. This leaflet may be handed to the patient directly when they come for appointment at the Macular Service. Additionally, potential participants may be contacted by phone. Following the phone conversation, the information leaflet will be sent by mail to the potential participants that showed interest in the study. Potential participants will be given an opportunity to ask questions/discuss the study either remotely (by phone) or face-to-face before consent for anonymized data collection is sought. Participants who are willing to consent will be asked to complete a consent form or, if consent is taken remotely, a researcher will complete the consent form in the participant’s stead. Throughout we will stress that participation in the research project is entirely voluntary, and participants can withdraw their consent at any time without it affecting their medical care in any way.

### 8.3 CONFIDENTIALITY

The Chief Investigator will preserve the confidentiality of participants taking part in the study and will abide by the UK General Data Protection Regulation and Data Protection Act 2018.

### 8.4 INDEMNITY

The University of Liverpool holds Indemnity and insurance cover with Marsh UK LTD, which apply to this study.

### 8.5 SPONSOR

The University of Liverpool will act as Sponsor for this study. It is recognised that as an employee of the University the Chief Investigator has been delegated specific duties, as detailed in the Sponsorship Approval letter.

### 8.6 FUNDING

Details of the Doctoral Network funding:

Remi Hernandez is funded by UoL EPSRC Doctoral Network in AI for Future Digital Health AI CDT.

### 8.7 AUDITS

The study may be subject to inspection and audit by the University of Liverpool under their remit as sponsor and other regulatory bodies to ensure adherence to GCP and the UK Policy Framework for Health and Social Care Research (v3.2 10th October 2017).

## 9. STUDY MANAGEMENT

The day-to-day management of the study will be coordinated through Mr Remi Hernandez.

## 10. END OF STUDY

The study will be completed 48 months after the end of the recruitment phase. The end date for recruitment is 31^st^ December 2024.

The study will end when all data collection is completed and the analysis have been fully performed, and data has been locked. End of study declaration and final study report will be submitted to HRA/REC and sponsor notifying them of the conclusion of the study. All the data will be archived for 10 years after the end of the study in line with the sponsor’s standard operating procedure on archiving, see Section 11 for details.

Suspension or early termination of the study will take place when there are substantial changes of patient care pathways, that cannot be mitigated by modifications of the study protocol to achieve the expected objectives.

## 11. ARCHIVING

Data for this study will comprise:

- Inclusion criteria and consent
- Demographics
- Medical history
- Medication
- Adverse events
- Binary files (images)

All the information will be collected and stored in accordance with UK General Data Protection Regulation and Data Protection Act 2018., and UoL and LUHFT guidelines.

Personal identifiable data will be physically stored in dedicated data centres on LUHFT as appropriate. Patient personal information collected will be stored at LUHFT in a study file meeting LUHFT SOPs and stored securely in the Clinical Eye Research Centre. Image assessment and grading by clinical CoIs will take place within the LUHFT environment either in the CERC or in the Liverpool Ophthalmic Reading Centre. Pseudo-anonymised study data will be anonymised before being transferred to UoL for further image analysis by using an encrypted memory key.

Electronic CRF related data and image data will be stored on the secured network space. All systems are continuously monitored. Appropriate measures are automatically taken whenever an alert is issued. The physical hardware will consist of enterprise-grade servers, networking, and storage solutions from tier 1 vendors and trustworthy and stable GNU/GPL solutions. Frequent backups are performed using the best enterprise backup solutions at LUHFT as well as UoL, and are physically stored in a fire-proof safe. The backup strategy will be implemented to support hourly, daily, monthly and yearly retention.

At the end of the study, the entire dataset will be archived in a reusable format. Archives encompass all raw data, meta-data, transformed data, transformation operations, deviations, version history, and audit trails. The comprehensive dataset will be archived on the LUHFT network driver so that the care team may still use for patient care. The anonymised data at the University side will be securely archived on the University’s Liverpool Data Catalogue, and accessed by future studies with ethical approval as appropriate. The comprehensive archive will remain propriety of the sponsor and will be preserved during a minimum period of 10 years.

## 12. PUBLICATION POLICY

The results of the study may be published in summary form, but the personal identity of any individual participants will not be revealed.

## Data Availability

N/A

## GLOSSARY OF ABBREVIATIONS

HRA: Health Research Authority
REC: Research Ethics Committee
AI: Artificial intelligence
CNN: Convolutional Neural Network
OCT: Optical coherence tomography
OCTA: Optical coherence tomography angiography
nAMD: Neovascular age-related macular degeneration
MNV: Macular neovascularisation
anti-VEGF: anti-Vascular Endothelial Growth Factor
FA: Fluorescein Angiography
ICGA: Indocyanine Green Angiography

### 14. APPENDICES

N/A

